# Medical Marijuana Had No Impact on Amphetamine Prescribing in Medicaid

**DOI:** 10.1101/2024.01.27.24301085

**Authors:** Errien M. Williams, Mariam Camara, Jessica L. Goldhirsh, Brian J. Piper

## Abstract

Due to the uncertainty of the health effects medical marijuana poses, states differ in their medical marijuana laws (MML). Long-term effects of marijuana are found to be like attention-deficit-hyperactive disorder (ADHD). With amphetamines being prescribed to treat ADHD symptoms we hypothesized that amphetamine prescriptions would increase in states implementing MML. The number of amphetamine prescriptions filled quarterly for each state from 2006 to 2021 were calculated. States with MML and dispensaries opened before 2020 were examined and states with no MML laws were the control. Prism was utilized to visualize the data and conduct four t-tests between the pre and post of MML+ versus MML-states. Three MML+ states were excluded due to limited post-MML data. Among the remaining states, 31 were MML+ and 17 were MML-. No significant differences were found in amphetamine prescribing (p > 0.30). Medical marijuana legalization did not have a statistically significant impact on amphetamine prescribing in Medicaid patients during the analyzed period. Contrary to the hypothesis, the results revealed a non-significant decrease in prescriptions in MML+ states. Further research with recreational cannabis laws or with electronic health records is warranted.

## 1. Introduction

Medical marijuana is a controversial topic within political and medical fields [1]. With a history of uncertainty regarding its benefits and its negative effects [2, 3], several states throughout America have yet to espouse unanimity concerning medical marijuana laws (MML). While many states have passed and implemented laws, others believe this matter to be immutable. One study found that long-term cannabis use impaired cognitive function to varying degrees, which may include emotional control, problem-solving abilities, and executive functioning [4]. Another longitudinal investigation concluded that marijuana use over time impacts short term learning, attention, and inhibition [5]. Many of the effects have similar characteristics to attention-deficit/hyperactivity disorder (ADHD), including inattention, forgetfulness, hyperactivity, and deficits in executive cognitive functioning [6, 7]. ADHD affects 5% of children, and 2.5% of adults worldwide [8]. ADHD is treated in the U.S. with amphetamines which increase both dopamine and norepinephrine at the synapse [9]. An increase in these neurotransmitters leads to improvements in executive function and overall reduction of ADHD symptoms [10].

Numerous reports have determined that ADHD is linked to an increased risk of substance use disorder (SUD) [8, 11, 12]. Cannabis is the most commonly used illicit drug among those with ADHD [8]. Further, ADHD can be associated with an increased lifetime use of cannabis use specifically [8]. The risk of marijuana use for ADHD patients is high and can be correlated with an increased risk of using other substances. In turn, the use of other substances has the potential to increase further ADHD symptoms [13]. Due to these findings [8, 11, 12, 13], further research is necessary to determine the relationship between MML and amphetamine prescriptions. We hypothesized that amphetamine prescriptions would increase after dispensaries opened in states where MML was implemented. Specifically, we used Medicaid data to assess the number of amphetamine prescriptions when corrected for the number of Medicaid and Children’s Health Insurance Program (CHIP) enrollees.

## 2. Results

Three states (Utah, Missouri, and Virginia) were excluded due to not meeting inclusion criteria. Thirty-one states including Washington D.C. met inclusion criteria and were categorized as MML+ and 17 states were MML-. No relationship was found between pre MML+ and post MML+ states (P = 0.2993) nor was there one for pre MML- and post MML-states (P = 0.9242, Figure 1). There was also no relationship found in pre MML+ vs pre MML-states (P = 0.3956) and post MML+ vs post MML– states (P= 0.9302, Figure 1). From pre to post in MML– states, there was a -16.5% decline in the slope while pre to post for MML+ states had a -64.0% reduction. Overall, there was no statistically significant difference between MML and amphetamine prescribing from 2006 to 2021 between states with or without MML.

**Figure 1.**
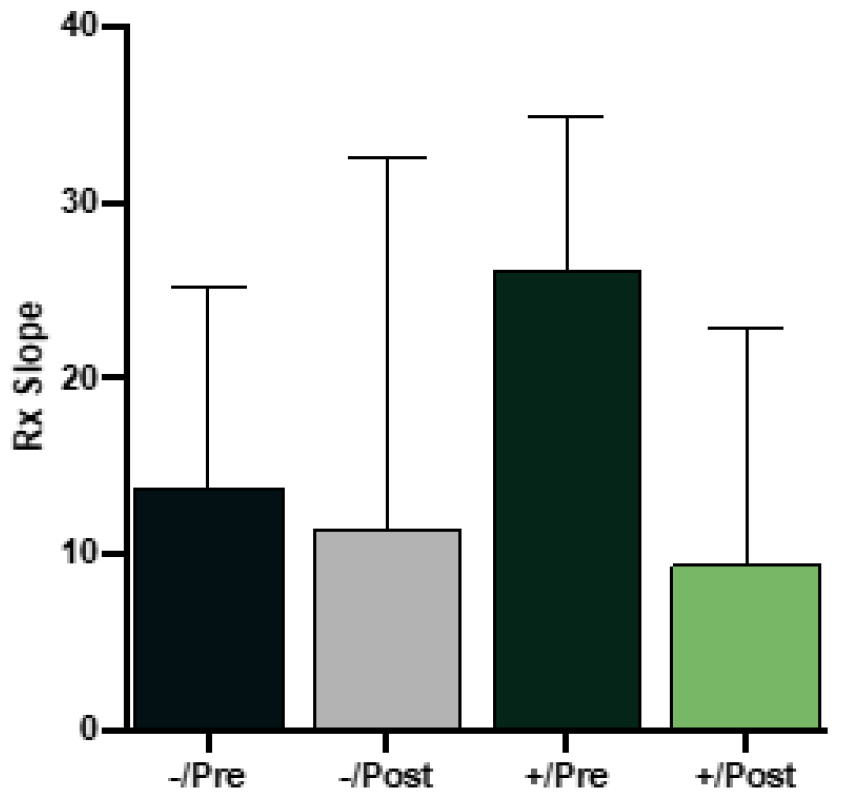
Slope of the amphetamine prescriptions (Rx) per 100,000 Medicaid/Children’s Health Insurance Program (CHIP) enrollees over time for states with (+) and without (–) medical marijuana laws implemented.

Further examination was completed on the change in slope (Rx over time) for the pre to post periods (Figure 2). Two MML-states, Georgia and Kentucky, had an increase (> +100) and two, Wisconsin and Wyoming, had a decrease (< -100). Similarly, among MML+ states, two increased, New Hampshire and New Mexico, and two decreased, Louisiana and Maine.

**Figure 2.**
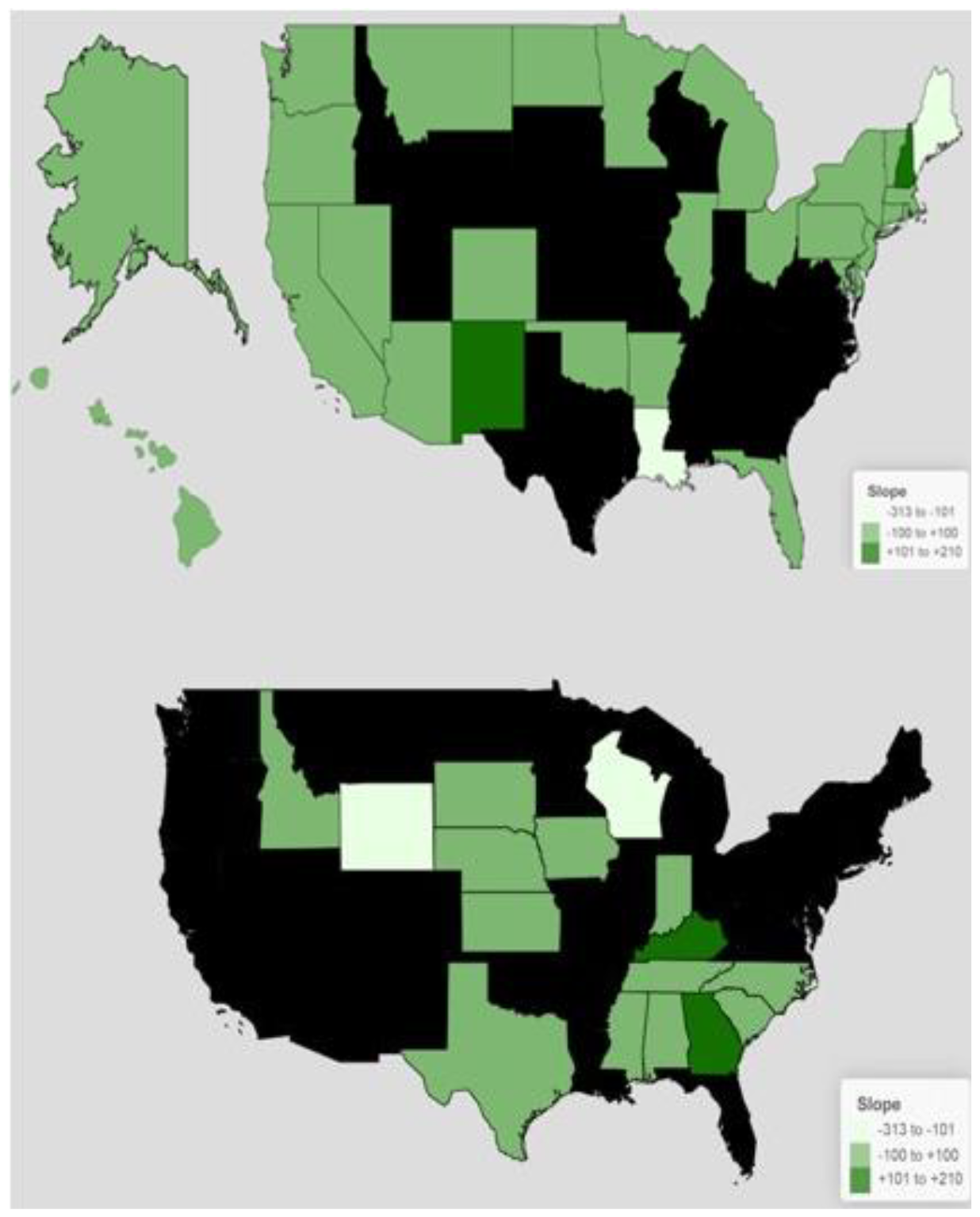
Heat maps demonstrating magnitude of prescription changes over time in amphetamines to Medicaid/Children’s Health Insurance Program patients in states with (top) and without (bottom) medical marijuana laws.

## 3. Discussion

This quasi-experimental report extends upon past research [19] and found that MML implementation and dispensary openings did not significantly impact amphetamine prescribing. In addition, the hypothesized finding was of an overall increase in prescribing in states which have active medical marijuana dispensaries. However, contrary to our hypothesis, there was evidence for an average decrease (−64.0%) in prescription slopes over time in states which have dispensaries. In a similar study, researchers tested overall prescriptions for Medicaid enrollees in areas with MML to determine if prescribing would decrease due to the availability of medical marijuana. They found a decrease in the prescriptions for pain, depression, nausea, psychosis, and seizures [20]. Another study with similar parameters was conducted and dealt with cannabis use and its effect on opioid and prescription drug consumption. Throughout the study, a slight decrease in this consumption was evident in locations with marijuana readily available, but there was no significant correlation seen between marijuana and the use opioids and prescription drugs [21].

This report also extends upon prior studies with a Drug Enforcement Administration database which has identified pronounced increases in amphetamine and lisdexamfetamine distribution over time [22, 23]. The mean slopes in Figure 1 were all greater than zero which indicates that amphetamine prescribing, when corrected for the number of enrollees, were increasing. Examination of the pre to post change in slopes (Figure 2) determined that only two MML- and two MML+ states had decreases (< -100) which again shows the MML had no impact on prescribing at a population level.

There were several limitations and caveats which should be considered. The potential neurocognitive effects with use of marijuana develop over a long period of time [5]. Studies have linked this long-term use to problems with controlling emotions, executive functioning, and even problem solving [4]. Other studies have linked long term use to problems with attention and short-term learning [5,6]. These potential symptoms mirror patients with ADHD as attentiveness, hyperactivity and problems with executive functioning are all linked to this disorder as well [6]. A prior report with methadone revealed that four states (Wisconsin, Tennessee, Oregon, and Vermont) accounted for almost two-thirds (64.0%) of opioid use disorder prescribing nationally which seems implausible [24]. We cannot discount the possibility of errors (e.g. dual data uploads) in the database [15] so, perhaps, these findings should be verified among those with private insurance. We can not exclude the possibility that some, and perhaps many, medical marijuana patients were previously using marijuana illegally. Future examination of the impact of recreational marijuana policies on continued escalation in amphetamine prescribing [Figure 1, 22, 23] should be conducted.

## 4. Methods

### 4.1 Data Sources

Data for the interrupted time series were collected from the Medicaid State Drug Utilization Data (SDUD) from 2006 to 2021 [Supplemental Table 1]. These data were reported by each state and included information on the drug name, number of prescriptions filled, and the National Drug Code (NDC) on a quarterly basis [14]. The sum of amphetamine prescriptions filled for each state’s quarter, including Washington D.C., for every year was calculated.

The number of Medicaid/CHIP enrollees was acquired using Medicaid and the Kaiser Family Foundation (KFF) [Supplemental Table 1] because the Centers for Medicare & Medicaid Services (CMS) began collecting states enrollment data using the Medicaid Budget and Expenditure System (MBES) in 2014 [15]. We used Medicaid to obtain enrollee data from June 2019 to June 2021. Then for the remaining years, 2006 to 2018, two KFF sources were used to obtain the total number of Medicaid/CHIP enrollees for all states including Washington DC [16, 17]. The enrollee data were recorded for the month of June for each year as the KFF source from 2006 to 2013 only included the mid-year values.

### 4.2 Procedures

States were categorized based on presence of a MML and when their first medical marijuana dispensary opened [18]. The dispensary opening date was included per state due to vast differences in the possible lag between the legalization of medical marijuana and its implementation. The states categorized as MML+ had MML and a dispensary open on or prior to 2006 through 2019. Three states with MML and a dispensary opened after 2019 were excluded (Missouri, Utah, Virginia) from the study because our collected SDUD cutoff was 2021. Lastly, states that were MML-served as the comparison. For each state, we divided the sum of prescriptions filled per quarter for each year by the number of Medicaid/CHIP enrollees for the year to determine the prescriptions filled per enrollee. Next, we created pre and post linear regression models for MML+ and MML– states. For MML+ states, we plotted twelve quarters pre and post the opening of a medical dispensary for all states except those which occurred in 2019. Those states only had eight quarters plotted pre and post dispensary opening due to our collected SDUD stopping at 2021.

For MML-states, we determined the date medical marijuana dispensaries opened in states with MML and utilized this as the break-point for the linear regression models [19]. For that calculation, we used the equation 20XX + (x /365.25), with x representing the total number of calendar days a dispensary opened and 365.25 accounting for leap years. There were five states (ME, MT, NV, OR, and WA) that reported only the month and year that dispensaries opened, so we assigned them the 15th of the month. The average was calculated to get the estimated date for states with no MML. We proceeded the same way as done for the states with MML in creating the pre and post linear regression models for states with no MML with 12 quarters being used. The resulting values were then adjusted for 100,000 enrollees for the MML+ and MML– states.

### 4.3 Statistical Analyses

The mean and standard error of the mean (SEM) were calculated for the slopes of prescriptions over time of the pre and post of MML+ states and MML-states. A slope > 0 was interpreted as increasing prescribing and a slope < 0 as decreasing prescribing. The change in slope (pre vs post) was also determined. Those values were then input in GraphPad Prism. Prism was also used to conduct four t-tests: between 1) pre + vs post + MML, 2) pre – vs post – MML, 3) pre + vs pre – MML, and 4) post + vs post – MML.

## Data Availability

All data produced in the present study are available upon request to the authors.

## Author Contributions

Conceptualization, J.L.G, E.M.W. and B.J.P..; methodology, E.M.W. and B.J.P.; software, E.M.W.; Formal analysis, E.M.W..; investigation, E.M.W., M.C.; resources, B.J.P.; data curation, ; writing—original draft preparation, E.M.W..; writing—review and editing, E.M.W., J.L.G., B.J.P..; visualization, E.M.W.; All authors have read and agreed to the published version of the manuscript.”

## Funding

This research received no external funding. Software was provided by NIEHS (T32 ES007060-31A1).

## Institutional Review Board Statement

The study was conducted in accordance with the Declaration of Helsinki, and approved by the Institutional Review Board of Geisinger (2021-0313 and approved 04-07-2021).

## Data Availability Statement

Data is available at <link>

## Acknowledgments

Thanks to Maria Tian, MBS for technical assistance.

## Conflicts of Interest

B.J.P. was (2019-21) part of an osteoarthritis research team supported by Pfizer and Eli Lilly. Other research is currently supported by the Pennsylvania Academic Clinical Research Center. Funders had no involvement in this study; in the collection, analyses, or interpretation of data; in the writing of the manuscript; or in the decision to publish the results. The other authors declare no conflict of interest.

## Notes

### Competing Interest Statement

The authors have declared no competing interest.

### Funding Statement

This study did not receive any funding.

